# Protocol for: mixed methods study on diversity of children with cochlear implants and their families engaging with the BEARS (Both Ears) virtual reality training games: improving clinical trial diversity and scale-up inclusiveness

**DOI:** 10.64898/2026.08.04.26359667

**Authors:** Helen Cullington, Sandra Driver, Ruth Nightingale, S. Sarah Somerset, Fleur Corbett, Marcus Jepson, Carmel Conefrey, Trishna Chauhan, Deborah Vickers

## Abstract

**Introduction:** We are currently working on the BEARS (Both Ears) virtual reality (VR) Randomised Controlled Trial. We are recruiting 272 deaf children and young people who use bilateral (both ears) cochlear implants (CI) to examine if using the BEARS VR games helps their hearing in background noise and ultimately their quality of life. Clinical trial participants should be representative of the population with the health condition, although this is rarely achieved in practice as people from ethnic minorities or those from more deprived areas may face barriers to participation.

**Study Design:** Mixed methods design

**Objectives:**

1. Use a literature review and collection of data from cochlear implant centres to establish the sociodemographic characteristics of deaf children aged 8 to 16 years with bilateral cochlear implants in the United Kingdom (UK).
2. a. Establish the sociodemographic characteristics of the families recruited to the BEARS clinical trial in the first six months, and compare with the age-matched population of deaf children with bilateral cochlear implants in the UK.

Analyse the BEARS clinical trial pre-screening diversity data.
Implement an established recruitment intervention method in a workshop to explore and optimise recruitment number and diversity.
Compare the final six months of recruitment data diversity with the initial six months.
3. Use interviews and focus groups to collect qualitative data from children and their families who chose not to take part in BEARS, clinicians, and family representatives (e.g. teachers) to explore barriers and facilitators to families taking part in BEARS.
4. Amend the BEARS scale-up plan based on new learning.

**Methods:** Literature and scoping reviews, quantitative analysis of BEARS recruitment data, in-depth interviews, paired interviews, focus groups

**Sample size:** Qualitative sample: 10-15 children aged 8-16 with bilateral CI, 10-15 parents/carers of children with bilateral CI, 10-12 clinicians and 10-12 family representatives.

**Significance:** This work will evaluate how diverse the BEARS clinical trial recruitment is and whether it is representative of the UK population of children with bilateral cochlear implants. We will investigate recruitment barriers and implement measures to try to improve recruitment diversity.

## Introduction

We are currently working on the BEARS (Both Ears) virtual reality (VR) Randomised Controlled Trial (RCT) (https://www.bears-ci.com/) funded by a National Institute for Health and Care Research (NIHR) programme grant (201608). We are recruiting 272 deaf children and young people aged 8 to 16 years who use bilateral (both ears) cochlear implants (CI). The trial was co-designed by young people with CIs and other stakeholders to address the listening difficulties the young deaf people told us they experienced. Children and young people with CIs have been involved throughout the design and refinement of the three BEARS VR training games and gave unique insights into what they wanted (1). Clinical trial participants are randomised to either use the BEARS VR training games at home, or to continue their usual care. We will evaluate if using the BEARS games helps participants’ hearing in background noise and ultimately their quality of life. If we find significant improvement, we plan to roll out the BEARS games to all deaf children using cochlear implants in the United Kingdom (UK); our trial ultimately aims to influence clinical practice and policy.

The proposed mixed methods development project will run in parallel with the programme grant. We aim to improve the quality and value of the BEARS clinical trial by ensuring recruitment is as diverse and representative as possible, and that our future plan to implement BEARS is inclusive. Our research seeks to serve all children with bilateral cochlear implants in the UK.

### Under-served groups in clinical research

Taking part in clinical research is generally a positive experience for patients (2) and improves the performance of healthcare organisations (3, 4). It is essential that people who take part in research reflect the population of those with the health condition (5). However, clinical trials often experience barriers to recruiting people from ethnic minority backgrounds (6) or people from deprived areas (7). A co-created National Health Service (NHS) report suggested that the primary reasons for under-representation in research are language barriers, accessibility and mistrust (8), and a health and social care research toolkit also highlighted stigma, communication and engagement barriers (9). Early work refining this protocol with our Public and Patient Involvement and Engagement (PPIE) group flagged similar barriers.

Deafness research is no exception to poor clinical trial diversity; reviews have shown that large study samples were not representative of the population of children with cochlear implants (10). Narrow inclusion criteria contribute to a lack of diversity. For example, the inclusion criteria for a study by Badarni-Zahalka et al (11) specified only children bilaterally implanted below the age of 3.5 years, with aided thresholds between 20 and 30 dBHL in both ears, word recognition scores above 75%, consistent device use, oral communication, mainstream education, and medium to high socioeconomic status. Hare et al (12) reported that standard research inclusion criteria for children with cochlear implants exclude approximately 75% of the population under examination. Some researchers are starting to address this. For example Nix et al (13) specifically assessed the demographic characteristics of the adults with cochlear implants in their study and found them comparable to the patient population from which they were sampled.

Inclusive research will not be attained until all research studies report their participant demographics in line with the recommendation by the International Committee of Medical Journal Editors (14). Research published in high-impact medical journals rarely reports participant ethnicity or socioeconomic status, or acknowledges this under-reporting as a limitation (15). A review of deafness intervention studies in the United States over 20 years showed that many demographic variables were under-reported, with parental education, family income, and parental occupation variables being least reported (16). Pittman et al (17) found low reporting of race/ethnicity in hearing intervention studies. Meinhardt et al (18) assessed 644 cochlear implant articles on interventional clinical trials and found that reporting of sociodemographic data (aside from gender) was very low (race 1%, ethnicity 1%, socioeconomic status 2%).

Under-served groups will vary from one study to another; we need to know which groups are under-served in the BEARS clinical trial so we can find solutions to ensure equality of access to the trial. These findings will not only influence recruitment for the BEARS clinical trial but will also inform the BEARS scale-up and implementation strategy. Incorporating qualitative investigation of recruitment has been shown to improve recruitment significantly in RCTs (19, 20). We will use some elements of the Qualitative research Integrated within Trials (QuinteT) recruitment intervention for ongoing RCTs (QRI-Two) to understand and ameliorate recruitment diversity issues (21).

### The population we wish to serve

We would like any benefits realised in the BEARS trial to be relevant to the whole population of children and young people with bilateral cochlear implants in the UK. Currently we do not know the demographic profile of children with cochlear implants in the UK, or indeed deaf children. We cannot assume that deaf children have the same characteristics as all children in the UK or that there are no social disparities in access to bilateral cochlear implants in deaf children.

Several UK organisations produce peer-reviewed literature and policy documents with information about deaf children with cochlear implants. It is unclear how much of the demographic information for these children is reported in items such as the CRIDE (Consortium for Research in Deaf Education) report, postcode data from cochlear implant centres, and reports from the National Deaf Children’s Society (NDCS). Some demographic data is known. We do know that a significant proportion (16 – 37%) of deaf children in the UK live in a low-income home (22). We also know that around 40% of deaf children with cochlear implants have other disabilities (23, 24). There is a higher incidence of deafness in some minority communities in the UK (25, 26). Data from 1001 children with bilateral cochlear implants in the UK included children with 23 different home languages (27). Thirteen percent of deaf children in the UK have English as an additional spoken language at home (28); this figure was higher in children with cochlear implants in one London centre (28%) (29). Hare, Sear and Vickers (30) have shown that there has been a significant reduction in the socioeconomic status of the families of children receiving cochlear implants over a 20-year period in a different London centre and that the proportion of children from non-native English-speaking families increased from 24 to 67%. These changes are partly attributed to population migration as immigration has increased (31) and periods of recession.

A scoping review offers a suitable methodology to identify the scope and volume of the literature available (32). Without this information we are unable to determine whether a representative sample of the population is participating in the BEARS clinical trial.

### Why would certain groups be excluded in the BEARS trial?

We would like the BEARS clinical trial data to be representative of all children and young people with bilateral CIs in the UK. In order to be included in the results of the BEARS clinical trial, children and families need to be cared for at a trial site, identified in the pre-screening process, contacted by the clinical team, consent to take part, and remain in the study. We are using a leaky pipe analogy to consider where we may be losing access to certain groups along the recruitment pathway (Figure 1). Previous research has shown that recruitment to RCTs can be improved by careful monitoring of the recruitment pathway using the SEAR framework (Screening Eligibility, Approach, Randomised or not) (33).

**Figure 1.**
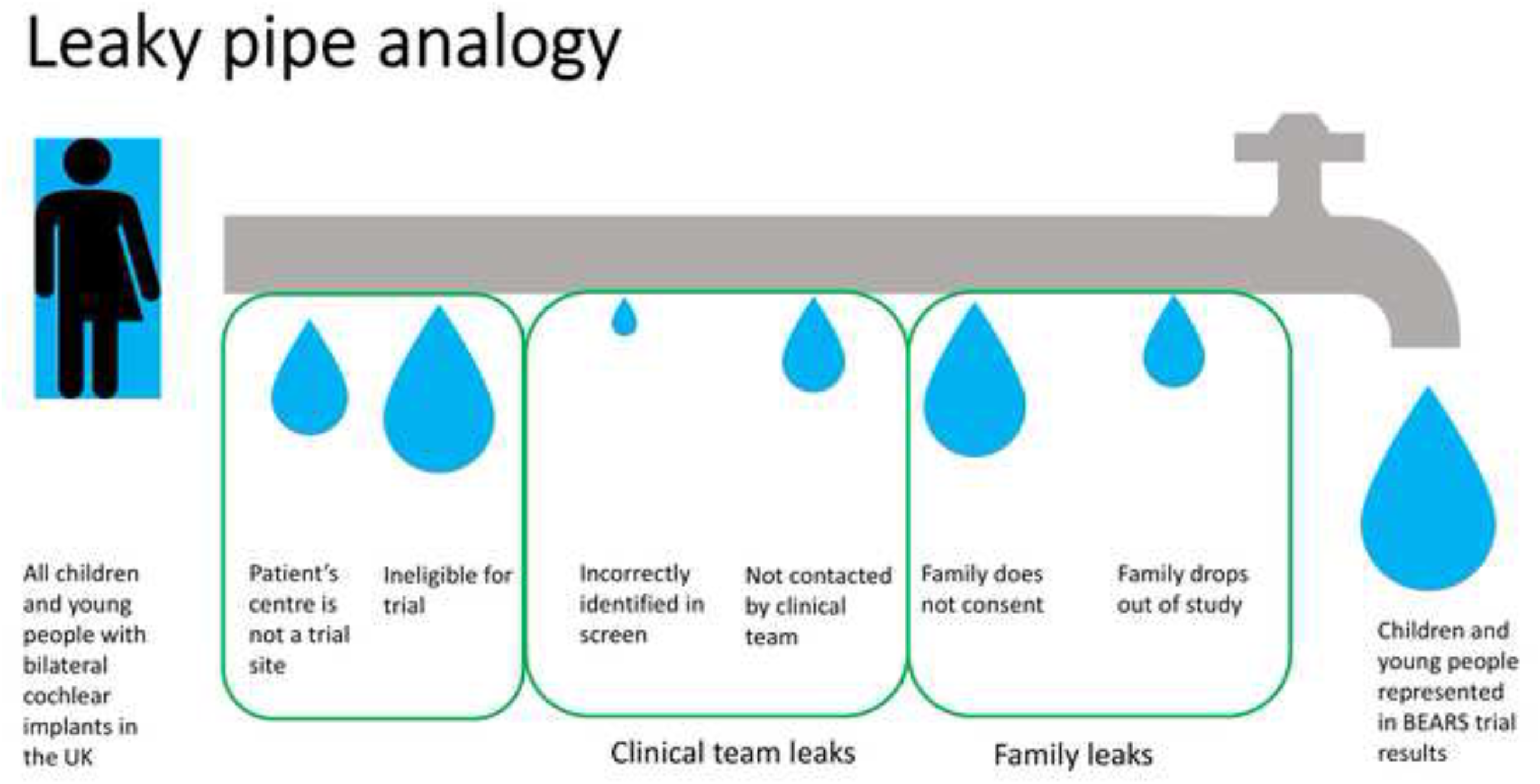
The leaky pipe analogy of people not being served in the BEARS clinical trial. This figure shows all UK children and young people (CYP) with bilateral cochlear implants on the left side, and the pipe analogy shows where CYP may drop out of the process, thus leaving a small proportion of CYP represented in the clinical trial results

### Which groups may be under-served in the BEARS trial?

The NIHR-INCLUDE project mentions the following examples of under-served groups that could be relevant to the BEARS clinical trial (34): different ethnic minority groups, educational disadvantage, people in full time employment (parents/carers), people in alternative residential circumstances (e.g. migrants, asylum seekers, care homes, prison populations, traveller communities, the homeless and those of no fixed abode), people living in remote areas, religious minorities, carers, people with language barriers, digital exclusion/disadvantage, people who do not attend regular medical appointments, looked after children, people with mental health conditions or multiple health conditions or physical disabilities.

Our target recruitment number for the BEARS clinical trial is 272 children spread across all UK paediatric cochlear implant centres. Due to the nature of participation and evaluation required for the programme grant trial, not all children and young people with bilateral cochlear implants can be included; inclusion and exclusion criteria are listed on the study website https://www.bears-ci.com/for-clinicians-2/.

The BEARS participant information sheets, consent forms, games, and questionnaires are all in English and translation of materials was outside the scope of the project. In addition, speech-in-noise tests have to be meticulously designed and validated in every language in order to cover all different speech sounds, so it is not a matter of just translation. Thus, one exclusion criterion for families is insufficient English for participation. As the participants are school-age children, it is likely that their English may be at a different level from their parents’, and with only one questionnaire for parents it is worth considering each family’s case individually where English is not the home language. We also exclude those with unstable outcomes from their implants, those who have not attended clinic appointments consistently, children who received cochlear implants older than 3 years (unless progressive hearing loss) and those with some additional needs. These criteria were necessary for us to answer the research questions and separate the effect of the BEARS games from other factors influencing outcomes, but we already suspect that recruitment is not representative.

Potential participants for the BEARS clinical trial will be identified through pre-screening measures at the implant centre where they are receiving usual care. The clinical team will review potential participants’ clinical notes, evaluating their implant history and demography. Following pre-screening, the implant centre research team should contact all those families considered eligible for the trial. However, we know that gate-keeping can occur whereby clinicians do not invite some patients to take part in research when they are eligible. This can often be with well-meaning intent, for example when the clinician feels the patient is too busy or that research participation would be too burdensome (35). However, this practice is not ethical and leads to skewed research results.

Even when contacted by the clinical care team, many families may choose not to participate in the trial. BEARS recruitment may not involve all genders equally; there is evidence to show that boys are more likely to play and spend more time on video games than girls (36). Our public engagement work at the Royal Institution Family Fun Day in February 2023 showed many more boys wanting to play the games than girls. Additionally, some families of children with cochlear implants have one or two deaf parents using British Sign Language. People who use sign language have much lower participation in research studies as they may be unable to access the study materials and may be unfamiliar with the concepts and terminology used in research projects (37). Unpublished data from a clinical trial of 323 UK adults with cochlear implants using an online intervention at home (Cullington) found that 47% of participants had Level 4 and above qualifications (degree, higher degree, professional qualifications). This is greater than that reported in the Census 2021 which found only 34% of people in England and Wales aged 16 and over with this qualification level. This suggests that people with cochlear implants choosing to take part in research are not representative in terms of qualification level.

### Are there specific barriers to participation related to the nature of the trial?

Virtual reality (VR) and active video games are a promising tool for rehabilitation but clinical adoption in general is low. A recent review of adults (within physical therapy) identified a gap in effective knowledge translation from clinicians to patients and the impact this has on VR uptake by participants (38). The BEARS trial uses VR training games, so it is important to understand the specific barriers and facilitators that participants with cochlear implants and their families may face related to using technology for home-based rehabilitation. The nature of the trial may also be another source of clinician gate-keeping: making assumptions about which families could use the equipment. A Meta Quest 2 virtual reality head-mounted display or an iPad is used for the BEARS games; these are standalone devices which do not require internet access or any additional home equipment. The family will also be lent headphones and a hand-held controller. The Meta Quest 2 and controller will need frequent recharging/battery change, which may be a barrier in homes struggling with energy costs.

The BEARS trial initially involved three clinic visits and then was reduced to two later in the trial, with other assessments completed online. Although some participant travel expenses will be paid, there are other barriers associated with travel for example time off work, family disruption and childcare. Unpublished data for people using cochlear implants at the University of Southampton Auditory Implant Service found that the median distance from home to the centre was 63 km, with 22% of patients living more than 100 km away.

## Materials and Methods

### Aims

We will evaluate the sociodemographic characteristics of families who have enrolled in the first six months of the BEARS clinical trial to establish whether we have a representative sample of the population of deaf children with cochlear implants in the UK. Literature and scoping reviews will help us to understand the sociodemographic characteristics of the population we wish to serve. Use of an established recruitment intervention method will attempt to improve the clinical trial recruitment; we will re-assess the sociodemographic characteristics of families in the final six months of the clinical trial to evaluate if we have improved diversity of participants. We will explore the barriers and facilitators to taking part in the BEARS clinical trial with stakeholders – focussing on people who declined participation. This new learning will improve the scale-up plan for the BEARS VR intervention.

### Setting

Data will be collected across all trial sites, which will be NHS or University cochlear implant centres in the UK.

### Design

Literature and scoping reviews; mixed methods design Four work packages (WP) will be completed corresponding to the study objectives.

### Work package 1

Research question 1: What are the sociodemographic characteristics of deaf children aged 8 to 16 years with bilateral cochlear implants in the United Kingdom?

#### Method

We will use the methodological framework proposed by Arksey and O’Malley (39) for scoping reviews, and report according to the PRISMA-ScR checklist (40). We will incorporate recommendations given by Levac et al (41) to clarify and enhance each stage, also incorporating quality appraisal of evidence. One key consideration is that the research question may evolve during the search process as the team become more familiar with the available evidence.

All children with cochlear implants in the UK are cared for by one of 17 paediatric cochlear implant centres. We will approach all centres to share anonymised data and may use a Freedom of Information request. We anticipate that they collect data about ethnicity, sex, home postcode etc; we would also like to know about family size, educational attainment, disability status and home language if possible. Other sources of data are charities, the Consortium for Research in Deaf Education, the UK National Registry of Hearing Implants, local authorities, and schools.

#### Inclusion criteria

- Any literature, reports, statistics, figures, studies, policies, documents, grey literature, articles etc which share any demographic characteristics of deaf children with CI
- Published in the last 10 years (2016 onwards)
- Data from the UK population

The output of this work package will be a summary of available evidence and whether it is valid and generalisable. If we find gaps in the available evidence, we will make recommendations for future reporting and highlight any areas of missing information.

### Work package 2

Research questions 2:

What are the sociodemographic characteristics of families recruited to the BEARS clinical trial in the first six months and children pre-screened for the BEARS trial? Are they representative of the age-matched population of deaf children with bilateral CIs in the UK? Does an established recruitment intervention method (QRI-Two) improve recruitment diversity?

#### Method

The first six months of BEARS recruitment sociodemographic data will be quantitatively analysed. The data we will have are:

- Age, sex, ethnicity, socioeconomic status as a proxy from home postcode (from BEARS screening appointment Case Report Form)
- Home language, parent/carer highest educational level and occupation, number of siblings (from BEARS baseline appointment Case Report Form)

We will then compare the sociodemographic profile of recruited children with each centre’s catchment area and the UK population to identify populations that are not being engaged. If available, we will also examine the sociodemographic profile of children pre-screened at each site. The data we will obtain from each centre will be group data describing the population they serve – ethnicity, sex, postcodes. If WP 1 reveals any quantitative data, that will also be used for comparison.

Data regarding the pre-screening of children with bilateral CI, eligibility for participation, and clinician contact with eligible families will be analysed for each trial site using the QuinteT SEAR framework (33) to determine 1. the percentage of children that the clinicians consider eligible for the BEARS clinical trial and reasons for ineligibility of others; 2. the percentage of eligible children and families contacted, and reasons why others were not contacted, who consented to participate, who declined to participate, reasons why declined if known, and who did not respond.

We will follow the QRI-Two protocol (21) to identify and estimate the scale of ‘hidden’ barriers in the recruitment strategy. These barriers may include inconsistencies in participant information, clinician discomfort or emotion related to beliefs in the effectiveness of the intervention, or any perceived conflict with their clinical roles(42). Depending on the above findings, rapid content analysis of BEARS trial documentation (patient information materials, consent forms, protocol) may be required; input from our PPIE group will help here. As knowledge about the sociodemographic profile of participants and recruitment process is generated, findings from the analysis of the initial recruitment data will be presented to the BEARS trial team, site clinicians, and PPIE lead at a workshop. We will work together to devise remedial actions and optimise the recruitment strategy in line with the INCLUDE recommendations (43), with the aim of improving recruitment within the remaining timeframe of the clinical trial (recruitment ends 31 July 2026). The INCLUDE recommendations are to ensure 1. eligibility criteria and recruitment pathway do not limit participation in unintended ways, 2. trial materials are developed with inclusion in mind, 3. trial staff are culturally competent, and 4. trusting partnerships with community organisations that work with ethnic minority groups.

We may also need to 1. change aspects of the protocol (for example, changing the times of research sessions may facilitate inclusion for some families), 2. change study materials e.g., advertising, and 3. educate and support clinicians. For example, if we find clinician gate-keeping has occurred, we will use education methods to address and decrease this (35).

We will make full use of the toolkit for Increasing participation of black Asian and minority ethnic (BAME) groups in health and social care research (9). The final stage of WP 2 will occur in October 2026, after BEARS recruitment ends. We will quantitatively analyse the final six months of recruitment data (February – July 2026, after the workshop) to assess whether the actions of WP 2 improved recruitment diversity.

### Work package 3

Research question 3: What are the barriers and facilitators to families taking part in the BEARS clinical trial?

#### Method

WP 3 will focus on children and families who declined participation in the BEARS trial. We want to understand more about the lived experience of those declining participation and the reasons for their lack of engagement. We will use qualitative semi-structured interviews, paired interviews and focus groups with families of children with CI, clinicians and family representatives/stakeholders such as teachers and volunteers. We will engage with the PPIE group to understand how we can engage under-represented groups from the trial and implement this into future plans for BEARS scale-up. We also want to explore why children and/or families might not engage with the BEARS intervention. This WP focuses on the:

1. Barriers and facilitators to family participation in the trial
2. Barriers and facilitators to the use of technology (i.e. using BEARS at home). If BEARS becomes part of routine care, what would support them to use it (scale-up)

We will ask clinicians what underpins their decision-making around the pre-screening process, who they invite to participate, their perceptions of why families decline to take part, and how this could impact scale-up. We will ask family representatives their perceptions of why families decline to take part in research, the use of technology such as a VR rehabilitation tool for children, and how this might impact scale-up.

Recruitment for WP3 began on 18 September 2024 when the ethics amendment was approved and continues until 31 July 2026.

#### Theoretical framework

Scaling up and implementing BEARS in clinical practice will require changes in behaviour at individual (e.g. child, parent, clinician), collective and organisational levels. Therefore, the Theoretical Domains Framework (TDF), a synthesis of 33 behaviour change theories into 14 domains (44), will inform WP3.

Using the TDF will enable us to conceptualise the barriers and facilitators to change, and together with findings from the process evaluation (conducted in the BEARS trial), will inform WP4 (45).

#### Participants and setting

Inclusion criteria:

- Children/young people with cochlear implants and their parents who are eligible for the BEARS trial but declined to take part
- Clinicians/gate-keepers (e.g. research audiologists, rehabilitationists) who are responsible for pre-screening and decision-making about whether child/family 1) meets the inclusion criteria, and 2) is subsequently invited to take part in the trial
- Family representatives/stakeholders from across the UK (e.g. teachers of the deaf, members of the voluntary sector such as ambassadors from the NDCS, the Cochlear Implanted Children’s Support group (CICS), and regional deaf societies).

#### Sampling and recruitment

Purposive sampling will be used to achieve maximum variation among participants in relation to:

1. children’s age, sex, ethnicity, study site, home language and family socioeconomic status
2. clinicians’ role, study site and research experience
3. family representatives’ role, organisation and area of the UK

We aim to recruit 10-15 children aged 8-16 with CI, 10-15 parents/carers, 10-12 clinicians and 10-12 family representatives. The final sample size will be determined by reaching data saturation. Children, parents and clinicians will be recruited from the study sites that are taking part in the BEARS trial. If necessary, we will use social media (e.g. British Cochlear Implant Group (BCIG), NDCS, CICS) to recruit families who declined to take part in the trial. Family representatives will be recruited via various methods including through the British Association of Teachers of Deaf Children and Young People (BATOD) network, CICS, NDCS, BCIG and social media. The letter of invitation for the BEARS project is amended to include text asking the family to say why they do not want to take part (either by email, phone, QR code/link to web questions on a Microsoft Form). The family can include their email address/phone number if they want to take part in an interview; otherwise, their reasons for not taking part remain anonymous.

#### Data collection

A mixture of qualitative methods including in-depth semi-structured individual interviews, paired interviews and focus groups will be used, as appropriate to each participant. Data collection will take place either virtually/online via videoconference, or in person depending on participants’ preferences. Interviews/focus groups will be recorded with consent/assent from the adults/children. The ethics amendment included verbal consent at the start of the interview; we do not want to introduce additional steps that may put people off taking part in the research, especially as these are families who have already declined participation in said no to the BEARS trial. We will make the study information as simple and accessible as possible and will translate where required. We will make the study information as simple and accessible as possible and will translate where required. If interpreters are required, we will provide.

Interviews are expected to last between 30-60 minutes and focus groups between 60-90 minutes. All interviews/focus groups will be transcribed by a General Data Protection Regulation (GDPR) compliant transcription service. Interviews will be guided by a topic guide informed by the TDF and developed with input from the PPIE group. All children taking part in an interview will be offered a £10 voucher.

#### Data analysis

Data will be analysed thematically using the Framework approach (46). This matrix-based analytic method is flexible and systematic and facilitates rigorous and transparent data management. The domains and constructs of the TDF framework will inform our analysis and interpretation of the data.

### Work package 4. Amend the BEARS scale-up plan based on new learning

Research question 4: How can we use this learning to improve the scale-up plan for the roll out of BEARS?

Preliminary data from the BEARS trial process evaluation will be used to directly complement the findings from this project to enable iterative refinement of the scale-up and implementation strategy.

The scale-up and implementation strategy seeks to optimise the trial approach for NHS adaptation of the BEARS games as a patient self-managed technology-based care option, ensuring equity and inclusivity.

This strategy is subject to iterative improvement from new learning and informed by Normalisation Process Theory (NPT) (47, 48), which comprises four key concepts:

1. Coherence: how clinicians and patients understand BEARS
2. Cognitive participation: buy-in and support for BEARS
3. Collective action: operational work required for BEARS
4. Reflexive monitoring: how BEARS is appraised and how work can be configured to enable it

We will also incorporate the programme grant’s early findings from the Normalisation MeAsure Development (NoMAD) questionnaire (49). A significant part of the latter months of the main BEARS trial focusses on implementation and roll out. The new learning on diversity from this project will augment the approach we are already taking.

We will continue to involve our PPIE group and will obtain feedback from them and other stakeholders as the scale-up plan evolves.

## Ethical considerations

The BEARS clinical trial was reviewed by the Yorkshire & The Humber - Sheffield Research Ethics Committee (Integrated Research Application System (IRAS) project ID 319903; REC reference 23/YH/0046) and the original approval was obtained on 14 March 2023. We submitted an amendment to the programme grant ethics (substantial amendment SA03) to include this development work; it was approved on 18 September 2024 (protocol 4.0 22 August 2024) by Yorkshire & The Humber - Sheffield Research Ethics Committee (IRAS project ID 319903; REC reference 23/YH/0046). The current approved protocol of the BEARS clinical trial is 7.0 14 July 2026. Ethics amendments for this study:

- Change to the invitation letter asking if parents/carers/children and young people who say no to BEARS would say their reasons on an online form and/or be willing to be contacted to explain their reasons
- To recruit parents/carers/children and young people/clinicians/family representatives to take part in interview/focus group (additional participant information sheets and consent/assent forms were submitted)
- To use already-collected BEARS data for additional analysis
- To collect demographic data on the pre-screening log
- To collect each site’s pre-screening log without participant identifiable

## Data sharing

The anonymized, group-level datasets generated for this study will be made publicly available in an open repository at the time of publication. Individual-level data cannot be shared due to ethical restrictions designed to protect the confidentiality of paediatric participants and their families. The minimal dataset required to replicate the findings will be provided, and any additional access requests for ethically restricted materials will be reviewed by the study’s institutional ethics committee. The qualitative data in WP3 will not be open access or put in a data repository after the project as this was not made explicit in the Participant Information Sheet. Therefore participants, in particular children/parents, have not consented to this.

## Patient and public involvement and engagement (PPIE)

We continue to have access to the established BEARS PPIE group. Since this development work focuses on equality, diversity and inclusivity, we have set up a new PPIE group covering a wide age range and including people with lived experience of deafness, cochlear implants and identifying as part of a minority community. The PPIE lead is author Trishna Chauhan.

Members of the PPIE group will be asked for their input to study documentation (consent/assent form, information sheet). As we are committed to engaging under-served groups, it will be particularly important that documents are accessible and not off-putting to potential participants. PPIE input will also be sought for the recruitment intervention analysis in WP 2, planning the interview questions in WP 3, the scale-up plan (WP 4), and dissemination (particularly to patients).

We are committed to maintaining a feedback cycle between the research team and the PPIE group to develop relationships, increase confidence and motivation. The lead author, Helen Cullington will take responsibility for providing feedback, either via the PPIE lead or directly to the PPIE contributor. HC will also keep a log of ’Did this lead to change?’ to track the impact of our PPIE.

## Status and timeline of the study

At the point of writing (July 2026), data collection for WP 1 and WP 2 are incomplete. The recruitment intervention workshop happened in October 2025. WP 3 focus groups have happened and analysis is ongoing, but responses to the invitation letter asking why families did not want to take part is still open. WP 4 is in progress but awaiting results from earlier WPs.

## Discussion

### Limitations of the study design

It may not be possible to ascertain the demographic profile of deaf children with cochlear implants in the UK from available data. The mandatory National Registry for Hearing Implants launched in mid 2025 so limited sociodemographic data will be available (ethnicity, social care involvement, home language). Participating BEARS sites may not have collected their pre-screening data in a consistent way that we can use. It is difficult to obtain data from busy NHS clinicians, especially those sites who are running the BEARS trial who may already be struggling to fit the BEARS research into their workload.

WP 3 sets out to obtain data from children and their families who chose not to take part in BEARS. Asking families who declined research involvement to be involved in research is challenging; however, we will meet with our PPIE group to generate ideas for recruiting these families.

### Dissemination plans

This project has diverse stakeholders - patients, the public, clinicians, teachers, funders, commissioners, managers, service leads, the NHS, hearing researchers, diversity researchers, cochlear implant companies, VR companies etc. We already have a communications plan for the BEARS programme grant with ideas of how to reach each sector. We have developed a new section on our website (https://www.bears-ci.com/diversity/) covering Diversity; we will keep this updated.

We will produce two peer-reviewed publications, presentations at conferences, and newsletter articles for the National Cochlear Implant Users Association (NCIUA) and Cochlear Implanted Children’s Support group (CICS). It has worked well for us previously to co-present with a patient; we will continue this for future conferences. We will produce an easy-read postcard to share our results with patients and the public. We will print 1000 and send bundles to all UK cochlear implant centres for their waiting rooms, for inclusion in NCIUA and CICS mailings, and our social media (LinkedIn BEARS Both Ears). We are particularly keen to disseminate the findings to the participants in this research and will ask them for contacts in their own communities where it may be appropriate to give out postcards. Our PPIE group will also be involved in dissemination. Including a diverse population in research is not just applicable to cochlear implants, or indeed deafness. We would like to share our learning more widely - to encourage other researchers to focus on research participant diversity. We will offer seminars for audiology teaching programs (both undergraduate and postgraduate) to influence a future generation of researchers.

## Data Availability

No datasets were generated or analysed during the current study. All relevant data from this study will be made available upon study completion.

## Acknowledgements

The authors thank the PPIE group members for their valuable contributions.

## Contributors

[HC, SD, RN, SS, FC, MJ, CC, TC, DV] conceived of the study and developed the protocol. [HC] wrote the first draft of the manuscript and subsequent revisions, with critical feedback from all other authors.

All authors contributed to the preparation of this manuscript and have reviewed and approved the final version.

## Competing interests

No potential conflicts of interest are reported by the authors.

This work is registered as part of the clinical trial that this work relates to:

1. ClinicalTrials.gov Clinical Trials Register, where it is identified as NCT05808543
2. ISRCTN Clinical Trials Register, where it is identified as ISRCTN92454702

